# Genetic variants in *DDX53* contribute to Autism Spectrum Disorder associated with the Xp22.11 locus

**DOI:** 10.1101/2023.12.21.23300383

**Authors:** Marcello Scala, Clarrisa A. Bradley, Jennifer L. Howe, Brett Trost, Nelson Bautista Salazar, Carole Shum, Miriam S. Reuter, Jeffrey R. MacDonald, Sangyoon Y. Ko, Paul W. Frankland, Leslie Granger, George Anadiotis, Verdiana Pullano, Alfredo Brusco, Roberto Keller, Sarah Parisotto, Helio F. Pedro, Laina Lusk, Pamela Pojomovsky McDonnell, Ingo Helbig, Sureni V. Mullegama, Undiagnosed Diseases Network, Emilie D. Douine, Bianca E. Russell, Stanley F. Nelson, Federico Zara, Stephen W. Scherer

## Abstract

Autism Spectrum Disorder (ASD) exhibits an ∼4:1 male-to-female sex bias and is characterized by early-onset impairment of social/communication skills, restricted interests, and stereotyped behaviors. Disruption of the Xp22.11 locus has been associated with ASD in males. This locus includes the three-exon *PTCHD1* gene, an adjacent multi-isoform long noncoding RNA (lncRNA) named *PTCHD1-AS* (spanning ∼1Mb), and a poorly characterized single-exon RNA helicase named *DDX53* that is intronic to *PTCHD1-AS*. While the relationship between *PTCHD1/PTCHD1-AS* and ASD is being studied, the role of *DDX53* has not been examined, in part because there is no apparent functional murine orthologue. Through clinical testing, here, we identified 6 males and 1 female with ASD from 6 unrelated families carrying rare, predicted-damaging or loss-of-function variants in *DDX53*. Then, we examined databases, including the Autism Speaks MSSNG and Simons Foundation Autism Research Initiative, as well as population controls. We identified 24 additional individuals with ASD harboring rare, damaging *DDX53* variations, including the same variants detected in two families from the original clinical analysis. In this extended cohort of 31 participants with ASD (28 male, 3 female), we identified 25 mostly maternally-inherited variations in *DDX53*, including 18 missense changes, 2 truncating variants, 2 in-frame variants, 2 deletions in the 3’ UTR and 1 copy number deletion. Our findings in humans support a direct link between *DDX53* and ASD, which will be important in clinical genetic testing. These same autism-related findings, coupled with the observation that a functional orthologous gene is not found in mouse, may also influence the design and interpretation of murine-modelling of ASD.

---

Autism Spectrum Disorder (ASD) refers to a group of neurodevelopmental conditions characterized by a heterogeneous early-onset impairment of social interaction and communication, combined with the presence of restricted, repetitive, and stereotyped behaviors and interests^1–4^. Although cognitive development is variable in affected individuals, these disorders are the leading contributor to disability in preschool-age children^3; 5; 6^. The underlying abnormalities in brain development and functional connectivity are presumed to be secondary to interactions between the genetic and environmental factors, making ASD a complex and multifactorial condition^2; 7^.

ASD can occur in association with pathogenic sequence-level variants and copy number variations (CNVs; also, structural variations, SVs) in specific neuronal genes^8–10^. In a recent comprehensive annotation of all publicly available ASD genome sequence data, 134 ASD-associated genes were identified and ∼15% of families studied had a clinically relevant rare variant. ASD-relevant gene lists are being curated by the Simons Foundation Autism Research Initiative (SFARI)^11^ and the Evaluation of Autism Gene Link Evidence (EAGLE)^12^ with both datasets available at https://gene.sfari.org.

Among the loci associated with ASD, chromosome Xp22.11 has been found to be disrupted in autistic males^13–16^. We present here a summary of the latest genotypic and phenotypic features mapping to this locus (Figure 1). It encompasses *DDX53*, *PTCHD1* (MIM * 300828), and the long noncoding RNA (lncRNA) *PTCHD1-AS* (*PTCHD1* Antisense RNA (Head-To-Head))^15^. *Ptchd1* mutant mice show deficient attention and cognition, while variants in the human *PTCHD1* gene are linked to intellectual disability, impaired vision and more rarely, autistic features (Autism X-linked 4, AUTSX4 - MIM # 300830)^17–19^. However, many deletions detected in autistic participants also disrupt exons of the upstream lncRNA *PTCHD1-AS* and/or the protein-coding gene *DDX53*^15; 18; 20^ leaving uncertainty of a strict genotype-phenotype correlation. In our previous work, deletion mapping of males with ASD indicated that the exon 2-4 region of *PTCHD1-AS* (encompassing *DDX53*) represents a critical region and mouse modeling reaffirms a role for the lncRNA (unpublished).

**Figure 1.**
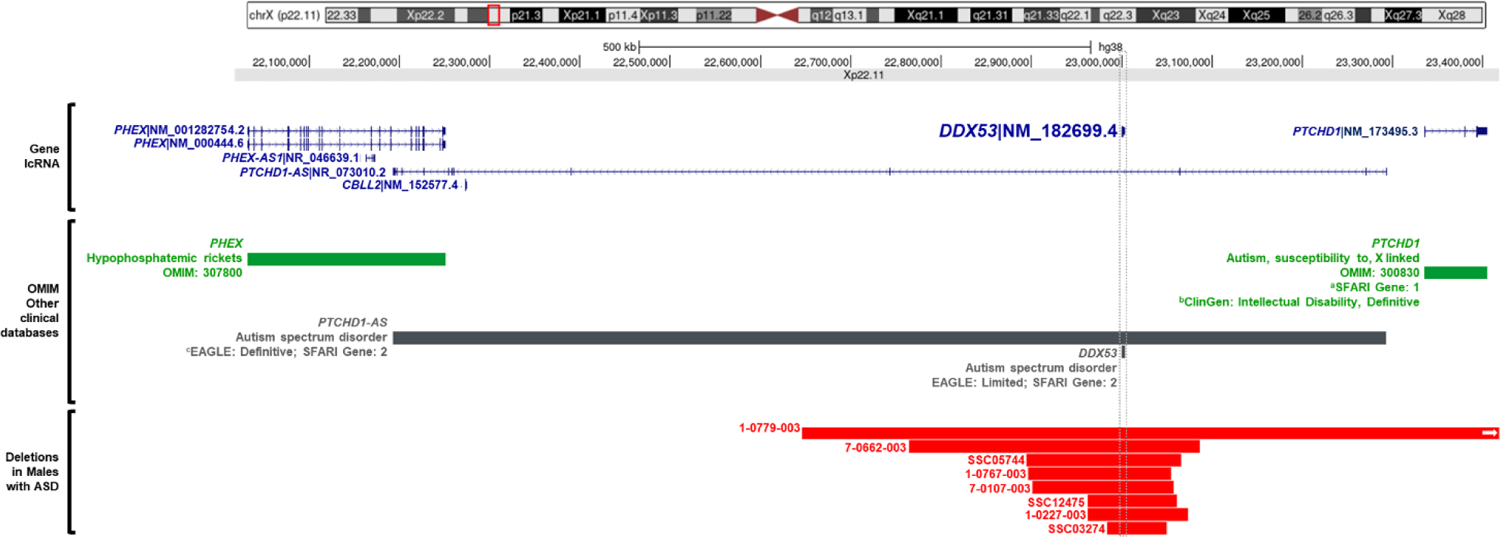
Summary of the genotypic and phenotypic features of the ASD-related Xp22.11 locus. The Xp22.11 locus includes *DDX53* (MIM * 301079), *PTCHD1* (MIM * 300828), and the long noncoding RNA (lncRNA) *PTCHD1-AS* (*PTCHD1* Antisense RNA (Head To Head)). A number of deletions identified in ASD participants were found to disrupt exons of the upstream lncRNA *PTCHD1-AS* and/or the *DDX53* gene. a = SFARI Gene with curated EAGLE gene scores: https://gene.sfari.org/; b = ClinGen https://clinicalgenome.org/; c = EAGLE gene curation https://www.ncbi.nlm.nih.gov/pmc/articles/PMC10357004.

*DDX53* is an intron-less gene of 3,630 nucleotides located within intron 3 of the canonical transcript of *PTCHD1-AS* encoding a 631-amino acid DEAD-box RNA helicase^21^. Phylogenetic analysis suggests *DDX53* and *DDX43* are highly related and that *DDX53* appears to be a retrogene derived from the 16-coding exon *DDX43* progenitor on chromosome 6^22^. The majority of retrogenes appear to originate from X-linked progenitors, possibly as a result of hypothesized compensation mechanisms^23; 24^. *DDX43* (72.8 kDa) and *DDX53* (71.2 kDa) proteins are also similar in size.

Moreover, *DDX53* is also described as being a cancer-associated gene (CAGE), since it was first detected in human gastric cancer cell lines^21^. It is overexpressed in human cancers, where it promotes cancer stem cell-like properties such as self-renewal, cell motility, tumor spheroid formation, and anticancer drug resistance^25; 26^. DEAD-box RNA helicases have crucial roles in mRNA metabolism, serving as ATP-dependent helicases and unwindases, chaperones, and mediators of association or dissociation with protein interactors (RNPases)^27; 28^. Additionally, RNA helicases act as transcriptional co-activators or co-repressors, driving target mRNAs either to protein synthesis or degradation^28^. As such, these enzymes contribute to coordinate gene expression programs within the cell^28^.

In this study, we investigated 7 participants (6 males and 1 female) with ASD, with or without impaired psychomotor development, in whom 6 distinct maternally-inherited variants in *DDX53* were identified. The study involving human participants was conducted according to the guidelines of the Declaration of Helsinki. Ethical review and approval were obtained locally through the Gaslini Children’s Hospital Research Committee for families I-VI (Comitato Etico della Regione Liguria, n.163/2018) and The Hospital for Sick Children Research Ethics Board. Family VI was consented at UCLA within IRB#15-000766 of the Undiagnosed Diseases Network. For all families, informed consent for the study and publication of research work was obtained from the participant or parents/ legal guardians of all the enrolled participants. After the identification of the index case, further individuals were included in the study cohort based on the identification of rare and potentially deleterious variants in *DDX53*, along with overlapping autistic and neurodevelopmental phenotypes. The initial participants were recruited through international collaboration involving clinical and research centers based in France, Italy, and the USA (further details available in the Supplementary Material), and also using GeneMatcher^29^. Enrolled participants were evaluated by pediatric neurologists, neuropsychiatrists, and neurogeneticists. For ASD assessment, thorough clinical examination and dedicated tests were employed, such as the Autism Diagnostic Interview-Revised (ADI-R) and Ritvo Autism Asperger Diagnostic Scale– Revised (RAADS–R) (Supplementary Material).

For genetic testing, trio-exome sequencing was performed for participant families I, II, and VI, and trio-whole genome sequencing was performed for VII. Participants #3, and #5 were investigated through the Next Generation Sequencing-based Autism/ID Xpanded Panel (GeneDx, Gaithersburg, USA) and participant #4 had targeted sequencing (GeneDx, Gaithersburg, USA) (Supplementary Material). Genomic DNA was extracted from peripheral blood lymphocytes of the probands and parents, with the sequencing being performed as described previously (Supplementary Material). Variants were filtered according to minor allele frequency ≤ 0.001 in Genome Aggregation Database (gnomAD), were assessed for levels of conservation (Genomic Evolutionary Rate Profiling – GERP), and functional impact was predicted according to different *in silico* tools (Supplementary Material), as well as the Variant Effect Predictor (VEP) pipeline from Ensembl.

Candidate variants in known disease genes were classified according to ACMG guidelines^30^. Variants were mapped onto the protein structures and intolerance to variation of the affected amino acid residues was analyzed using the Metadome online software (https://stuart.radboudumc.nl/metadome/method)^31^. Copy number variation (CNV) assessment was performed either through array comparative genomic hybridization (aCGH) or whole genome/exome-based CNV calling (Supplementary Material). The DECIPHER database and Database of Genomic Variants were used to interpret detected CNVs^32; 33^. *DDX53* variants are reported according to the NM_182699.4 transcript, corresponding to the unique isoform of the DDX53 protein NP_874358.2 (https://www.ncbi.nlm.nih.gov/gene/168400).

We identified six distinct variants in *DDX53* in the reported individuals: c.871A>G, p.(Arg291Gly) in #1; c.834G>A, p.(Met278Ile) in #2; c.567_568insTGCAGGT, p.(Glu190Cysfs*3) in #3 and #4; c.977G>A, p.(Ser326Asn) in #5; c.1736C>T, p.(Ala579Val) in #6; c.259G>T, p.(Gly87Trp) in #7. All variants were maternally inherited and segregated with the phenotypes in all families (Figure 2A). In family III, both participants harbored the same p.(Glu190Cysfs*3) variant, although no information about a potential non-skewed X-inactivation in the female sibling (#4) was available. In family IV, the p.(Ser326Asn) variant identified in the proband (#5) was mosaic (4%) in the mother. In family V, the brother of participant #6 was reported to show autistic features, but segregation analysis could not be performed. In family VI, there were no other maternal male relatives for further segregation analysis available. All of the detected *DDX53* variants are rare (maximum allele frequency 0.000005534) and absent in the hemizygous state in gnomAD. The missense variants p.(Gly87Trp), p.(Met278Ile), p.(Arg291Gly), p.(Ser326Asn), and p.(Ala579Val) affect conserved residues (GERP scores ranging between 3.55 and 4.59) within or in close proximity to the KH, DEXDc, and HELICc domains (Figure 2B).

**Figure 2.**
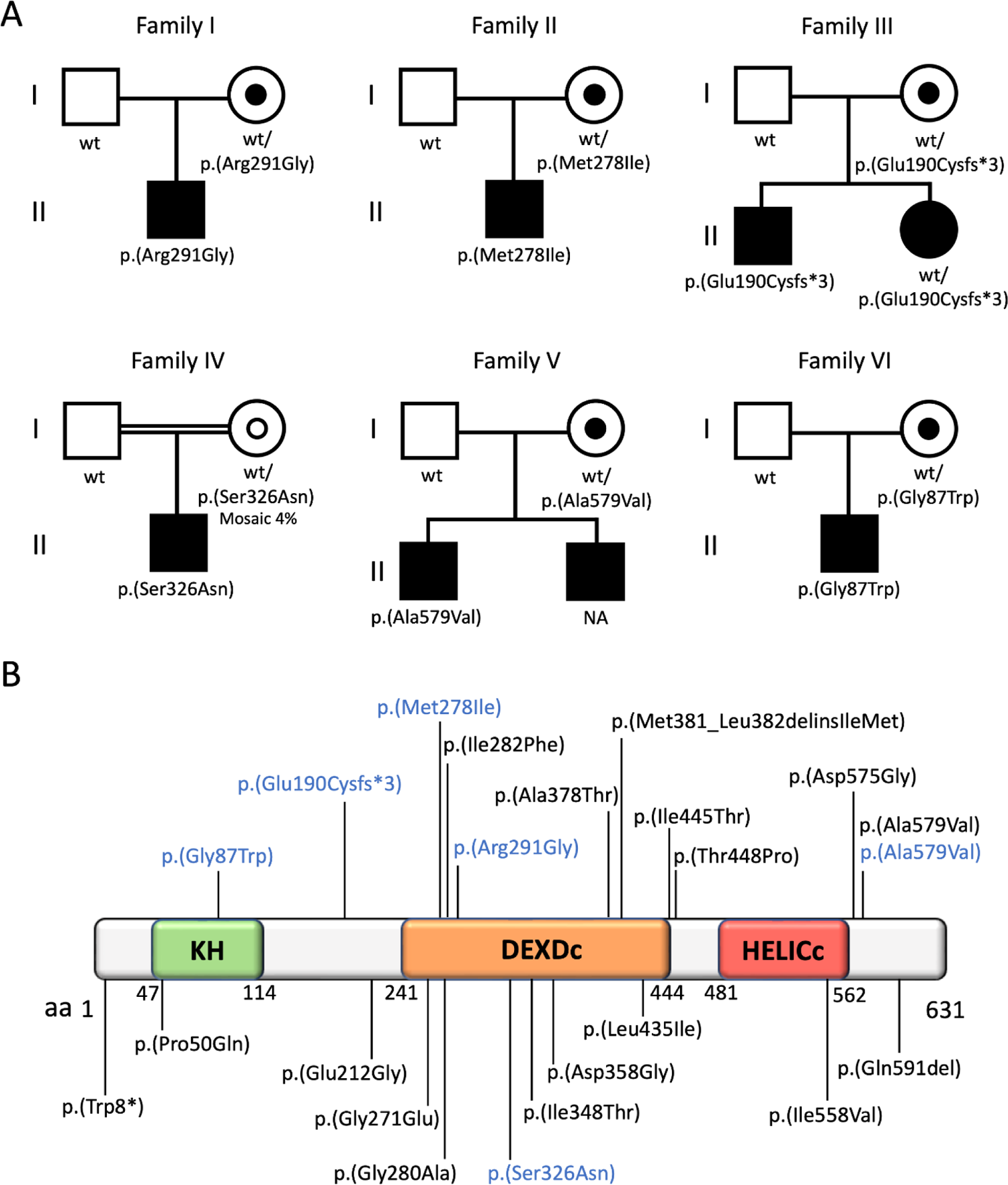
Pedigrees of the reported families and genetic findings in all *DDX53* participants. A) Pedigrees of the six families showing the segregation of the *DDX53* variants in affected individuals and the parents. All variants were inherited from unaffected mothers. The carrier status is indicated by small-filled circles. The mosaic status of the mother of the proband #5 is indicated by a small empty circle. **B)** Schematic representation of the total of the *DDX53* variants mapped to the unique protein isoform (NP_874358.2). The variants identified in the described families and those identified in participants from the SFARI and MSSNG databases are reported in blue and black, respectively. DEXDc = DEAD-like Helicases superfamily domain; HELICc = helicase conserved C-terminal domain; KH = K homology domain.

These changes are predicted to be damaging according to *in silico* tools (Table S1). The frameshift variant p.(Glu190Cysfs*3) is likely to lead to either nonsense-mediated mRNA decay (NMD) or the formation of a truncated transcript. Pathogenicity using the ACMG classifications could not be definitively assigned for this gene, as ACMG guidelines are only applicable for genes with already established gene-disease associations. The predicted effects of these variants are indicated in Tables 1 and 2.

**Table 1.**
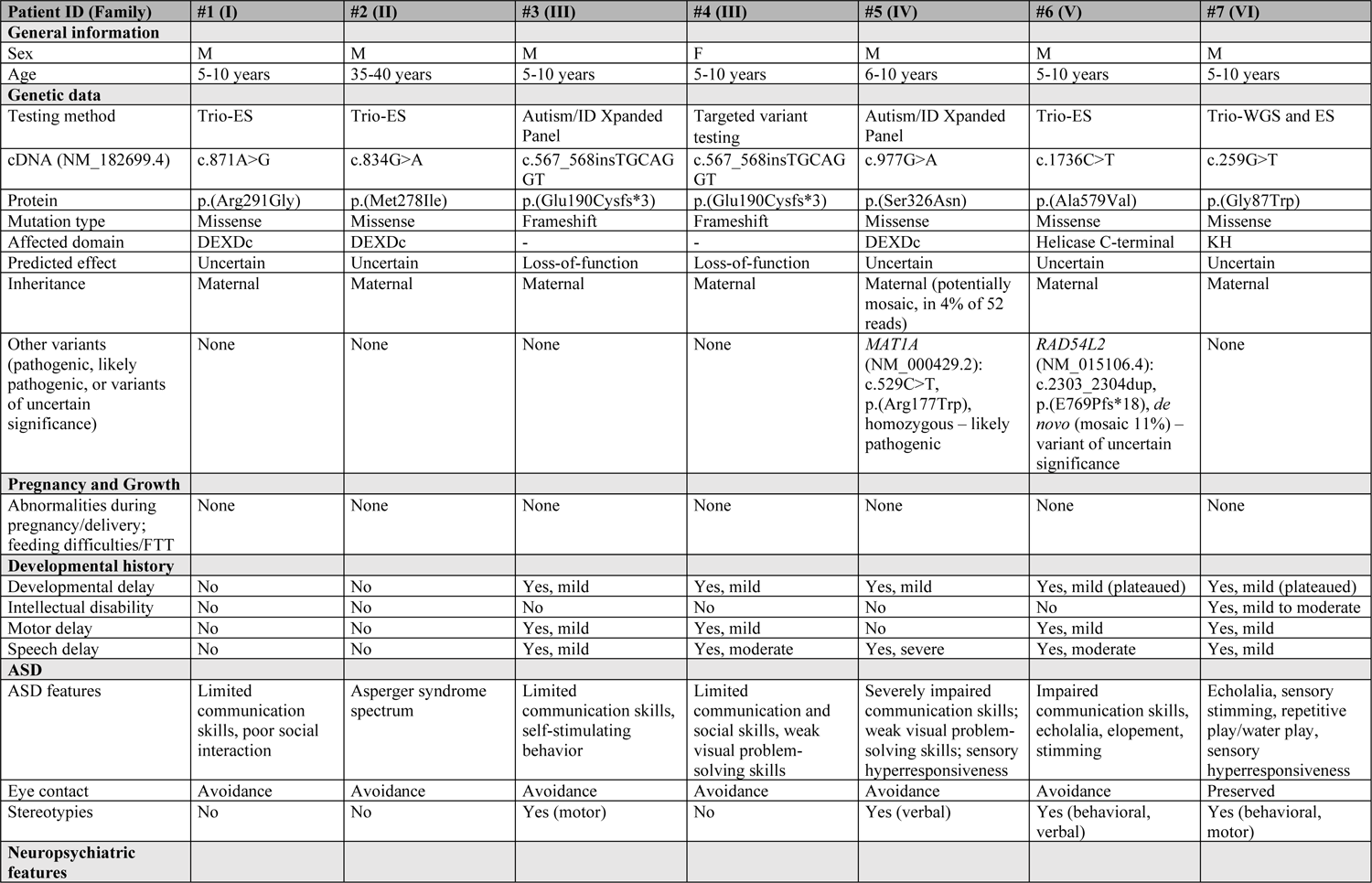
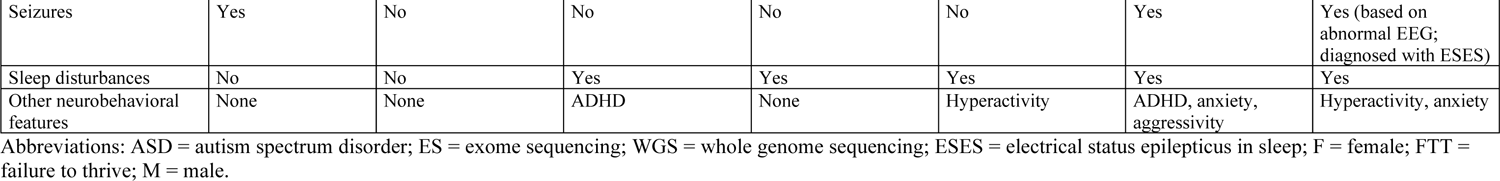
Clinical features of participants with *DDX53* variants.

| Patient ID (Family) | #1 (I) | #2 (II) | #3 (III) | #4 (III) | #5 (IV) | #6 (V) | #7 (VI) |
| --- | --- | --- | --- | --- | --- | --- | --- |
| **General information** |  |  |  |  |  |  |  |
| Sex | M | M | M | F | M | M | M |
| Age | 5-10 years | 35-40 years | 5-10 years | 5-10 years | 6-10 years | 5-10 years | 5-10 years |
| **Genetic data** |  |  |  |  |  |  |  |
| Testing method | Trio-ES | Trio-ES | Autism/ID Xpanded Panel | Targeted variant testing | Autism/ID Xpanded Panel | Trio-ES | Trio-WGS and ES |
| cDNA (NM_182699.4) | c.871A>G | c.834G>A | c.567_568insTGCAG GT | c.567_568insTGCAG GT | c.977G>A | c.1736C>T | c.259G>T |
| Protein | p.(Arg291Gly) | p.(Met278Ile) | p.(Glu190Cysfs*3) | p.(Glu190Cysfs*3) | p.(Ser326Asn) | p.(Ala579Val) | p.(Gly87Trp) |
| Mutation type | Missense | Missense | Frameshift | Frameshift | Missense | Missense | Missense |
| Affected domain | DEXDc | DEXDc | - | - | DEXDc | Helicase C-terminal | KH |
| Predicted effect | Uncertain | Uncertain | Loss-of-function | Loss-of-function | Uncertain | Uncertain | Uncertain |
| Inheritance | Maternal | Maternal | Maternal | Maternal | Maternal (potentially mosaic, in 4% of 52 reads) | Maternal | Maternal |
| Other variants (pathogenic, likely pathogenic, or variants of uncertain significance) | None | None | None | None | *MAT1A* (NM_000429.2): c.529C>T, p.(Arg177Trp), homozygous – likely pathogenic | *RAD54L2* (NM_015106.4): c.2303_2304dup, p.(E769Pfs*18), *de novo* (mosaic 11%) – variant of uncertain significance | None |
| **Pregnancy and Growth** |  |  |  |  |  |  |  |
| Abnormalities during pregnancy/delivery; feeding difficulties/FTT | None | None | None | None | None | None | None |
| **Developmental history** |  |  |  |  |  |  |  |
| Developmental delay | No | No | Yes, mild | Yes, mild | Yes, mild | Yes, mild (plateaued) | Yes, mild (plateaued) |
| Intellectual disability | No | No | No | No | No | No | Yes, mild to moderate |
| Motor delay | No | No | Yes, mild | Yes, mild | No | Yes, mild | Yes, mild |
| Speech delay | No | No | Yes, mild | Yes, moderate | Yes, severe | Yes, moderate | Yes, mild |
| **ASD** |  |  |  |  |  |  |  |
| ASD features | Limited communication skills, poor social interaction | Asperger syndrome spectrum | Limited communication skills, self-stimulating behavior | Limited communication and social skills, weak visual problem-solving skills | Severely impaired communication skills; weak visual problem-solving skills; sensory hyperresponsiveness | Impaired communication skills, echolalia, elopement, stimming | Echolalia, sensory stimming, repetitive play/water play, sensory hyperresponsiveness |
| Eye contact | Avoidance | Avoidance | Avoidance | Avoidance | Avoidance | Avoidance | Preserved |
| Stereotypies | No | No | Yes (motor) | No | Yes (verbal) | Yes (behavioral, verbal) | Yes (behavioral, motor) |
| **Neuropsychiatric features** |  |  |  |  |  |  |  |
| Seizures | Yes | No | No | No | No | Yes | Yes (based on abnormal EEG; diagnosed with ESES) |
| Sleep disturbances | No | No | Yes | Yes | Yes | Yes | Yes |
| Other neurobehavioral features | None | None | ADHD | None | Hyperactivity | ADHD, anxiety, aggressivity | Hyperactivity, anxiety |

Abbreviations: ASD = autism spectrum disorder; ES = exome sequencing; WGS = whole genome sequencing; ESES = electrical status epilepticus in sleep; F = female; FTT = failure to thrive; M = male.

**Table 2.** Genetic and clinical features of ASD participants with *DDX53* variants in the MSSNG and SFARI databases.

| Source | ID# | Sex | Variant (NM_182699.3); GRCh38 | Inheritance | Consequence | AF (gnomAD); Decipher | Hemizygous in gnomAD | *In silico* pathogenic impact (deleterious vs benign)* | Predicted effect | Affected *DDX53* domain | Phenotype | Other pathogenic variants |
| --- | --- | --- | --- | --- | --- | --- | --- | --- | --- | --- | --- | --- |
| SPARK | SP0003053 | F | c.24G>A, p.(Trp8*) | Unknown | Stopgain | 0.0000182 | 0 | High (4 vs 1) | Loss-of-function | - | ASD | - |
| MSSNG | AU2440301 | M | c.149C>A, p.(Pro50Gln) | Maternal | Missense | 0.00000892 | 0 | Medium (7 vs 11) | Uncertain | KH | ASD | - |
| SPARK | SP0039198† | M | c.567_568insTGCAGGT, p.(Glu190Cysfs*3) | Maternal | Frameshift | 0 | 0 | High (no benign) | Loss-of-function | - | ASD | - |
| SPARK | SP0042895† | F | c.567_568insTGCAGGT, p.(Glu190Cysfs*3) | Maternal | Frameshift | 0 | 0 | High (no benign) | Loss-of-function | - | ASD | - |
| SPARK | SP0035110 | M | c.635A>G, p.(Glu212Gly) | Maternal | Missense | 0 | 0 | Medium (8 vs 11) | Uncertain | - | ASD | - |
| SPARK | SP0137474 | M | c.812G>A, p.(Gly271Glu) | Maternal | Missense | 0.00000894 | 0 | High (16 vs 3) | Uncertain | DEXDc | ASD | - |
| SPARK | SP0032790 | M | c.839G>C, p.(Gly280Ala) | Maternal | Missense | 0.0000277 | 0 | Moderate (8 vs 10) | Uncertain | DEXDc | ASD | - |
| SPARK | SP0115897 | M | c.844A>T, p.(Ile282Phe) | Unknown | Missense | 0.0000563 | 0 | Medium (10 vs 9) | Uncertain | DEXDc | ASD | - |
| SPARK | SP0241833 | M | c.1043T>C, p.(Ile348Thr) | Maternal | Missense | 0 | 0 | High (14 vs 5) | Uncertain | DEXDc | ASD | - |
| SPARK | SP0030523 | M | c.1073A>G, p.(Asp358Gly) | Maternal | Missense | 0 | 0 | High (12 vs 7) | Uncertain | DEXDc | ASD | - |
| SPARK | SP0005644 | M | c.1132G>A, p.(Ala378Thr) | Unknown | Missense | 0 | 0 | High (12 vs 6) | Uncertain | DEXDc | ASD | - |
| SPARK | SP0052296 | M | c.1303C>A, p.(Leu435Ile) | Unknown | Missense | 0 | 0 | Medium (8 vs 11) | Uncertain | DEXDc | ASD | - |
| SSC | SSC10709 | M | c.1334T>C, p.(Ile445Thr) | Maternal | Missense | 0.00000892 | 0 | Medium (9 vs 9) | Uncertain | - | ASD | - |
| SPARK | SP0038972 | M | c.1342A>C, p.(Thr448Pro) | Maternal | Missense | 0 | 0 | Moderate (5 vs 13) | Uncertain | - | ASD | - |
| SSC | SSC10563 | M | c.1672A>G, p.(Ile558Val) | Maternal | Missense | 0 | 0 | Medium (7 vs 12) | Uncertain | HELICc | ASD | - |
| SPARK | SP0126624 | M | c.1724A>G, p.(Asp575Gly) | Maternal | Missense | 0.0000109 | 0 | Medium (7 vs 11) | Uncertain | C-terminal | ASD | - |
| MSSNG | 1-0551-003 | M | c.1724A>G, p.(Asp575Gly) | Maternal | Missense | 0.0000109 | 0 | Medium (7 vs 11) | Uncertain | C-terminal | ASD | - |
| SPARK | SP0090623‡ | M | c.1736C>T, p.(Ala579Val) | Unknown | Missense | 0.0000269 | 0 | Medium (6 vs 12) | Uncertain | C-terminal | ASD | - |
| SPARK | SP0066816 | M | c.1771_1773del, p.(Gln591del) | Unknown | Inframe deletion | 0 | 0 | High (no benign) | Uncertain | C-terminal | ASD | - |
| SPARK | SP0066817 | M | c.1771_1773del, p.(Gln591del) | Unknown | Inframe deletion | 0 | 0 | High (no benign) | Uncertain | C-terminal | ASD | - |
| MSSNG | MSSNG00067-003 | M | c.1143_1144delGCinsTA, p.(Met381_Leu382delinsIleMet) | Maternal | Inframe indel | 0 | 0 | Medium (1 vs 0) | Uncertain | DEXDc | ASD | *POGZ* (NM_015100.4) c.392dup, p.(Met132HisfsTer9), *de novo* |
| MSSNG | 1-0485-003 | F | Xp22.11 (22987677-23009507)x1 | Maternal | Deletion (21.8kb) | Absent in Decipher | NA | NA | Loss-of-function | - | ASD | chr7:70441029-70771836, deletion (including *AUTS2*), *de novo* |
| MSSNG | 7-0111-003 | M | Xp22.11 (23002082-23002458)x1 | Unknown | Deletion within the 3'UTR | Absent in Decipher | NA | NA | Uncertain | NA | ASD | - |
| MSSNG | 1-0627-003 | M | Xp22.11 (23002148-23002522)x1 | Maternal | Deletion within the 3'UTR | Absent in Decipher | NA | NA | Uncertain | NA | ASD, ADHD | - |
| MSSNG | 1-0627-005 | M | Xp22.11 (23002148-23002522)x1 | Maternal | Deletion within the 3'UTR | Absent in Decipher | NA | NA | Uncertain | NA | ASD, ADHD | - |
| MSSNG | **1-0627-006** | M | Xp22.11 (23002148-23002522)x1 | Maternal | Deletion within the 3'UTR | Absent in Decipher | NA | NA | Uncertain | NA | ASD, ADHD | - |
| MSSNG | **1-0627-007** | M | Xp22.11 (23002148-23002522)x1 | Maternal | Deletion within the 3'UTR | Absent in Decipher | NA | NA | Uncertain | NA | ASD, ADHD | chr16:83861601-85760600, deletion, *de novo* |

Abbreviations: ADHD = attention-deficit/hyperactivity disorder; ASD = autism spectrum disorder; F = female; M = male; NA = not available. Genome Build GRCh38. \* According to *in silico* tools predictions available in Varsome (https://varsome.com) † These patients correspond to subjects #3 (SP0039198) and #4 (SP0042895) from the main cohort, respectively. ‡ This patient (SP0090623) corresponds to subject #6 from the main cohort. ACMG guidelines could not be applied to determine pathogenicity of DDX53 variants as these should only be used for genes with established gene disease/disorder associations and not novel/potential disease/disorder genes.

The *DDX53* participants included in our cohort presented with marked ASD-related features, including poor social interaction, limited communication skills, self-stimulating behaviors, weak visual problem-solving skills, and sensory hyperresponsiveness (Table 1). Eye contact avoidance was observed in all cases but #7. Motor and/or verbal stereotypies were present in four participants (#3, #5, #6, and #7), sometimes in association with ritualistic behaviors (Table S2). A form of high-functioning autism within the spectrum of Asperger syndrome was diagnosed in #2. Interestingly, while two participants from Family III presented with similar overlapping ASD phenotypes, female participant #4 showed a more severe phenotype including significant speech limitation, social avoidance, sensory hypersensitivity, and impaired visual problem-solving skills (no additional variants explaining this were found). Autistic features were associated with impaired psychomotor development in five out of seven participants (#3-#7), variably affecting speech development and/or motor abilities. However, this developmental delay only led to variable skills, generally in the mild to moderate range, in participant #7. Additional neuropsychiatric manifestations consisted of seizures (#1, #6, and #7), heterogeneous sleep disorders (#3-#7), and behavioral comorbidities (#3, #5-#7), such as attention-deficit hyperactivity disorder (ADHD), anxiety, and aggressiveness.

After the identification of the *DDX53* variants in the clinical cohorts, we searched the SFARI (https://www.sfari.org/resource/sfari-gene/) and MSSNG (https://research.mss.ng) databases. Within SFARI^11; 34^, we screened for *DDX53* variants in the Simons Simplex Collection (SSC) and Simons Foundation Powering Autism Research (SPARK) datasets. SSC consists of WGS data from 2,600 simplex families, each containing one proband with ASD as well as unaffected parents and siblings. SPARK (WES1, WES2) contains the whole exome sequence for 43,259 participants (including 21,900 with ASD) with phenotype information. Additionally, we screened the MSSNG WGS database^8^; ^35–37^, which at the time consisted of 11,312 individuals, including 5,100 with ASD.

By screening the SSC, SPARK, and MSSNG cohorts, we detected 19 additional interesting variations in *DDX53* in 24 individuals, including 22 males and 2 females, presenting with mostly isolated ASD. These included 13 missense changes, 1 stopgain, 2 in-frame variants, 2 distinct deletions within the 3’UTR and one deletion CNV (Table 2). Fourteen of these variations were of maternal origin, whereas information about the segregation was not available in the remaining cases. Resembling those identified in our cohort, the *DDX53* variants detected in participants from the open-science autism datasets are exceptionally rare (max allele frequency of 0.0000563) and are absent in hemizygous state in gnomAD. Most missense changes affect conserved residues within different functional domains of the DDX53 helicase (Figure 2B) and are predicted deleterious by *in silico* tools (Table S3). The novel truncating variant p.(Trp8*) was predicted to exert a deleterious effect on protein expression, whereas the in-frame changes (p.(Gln591del) and p.(Met381_Leu382delinsIleMet) resulted in the deletion or substitution of highly conserved amino acids, respectively. Of note, in the SPARK dataset, we also identified participants that were already part of our main cohort: subjects #3 (SP0039198), #4 (SP0042895), and #6 (SP0090623).

Deletions within the 3’UTR of *DDX53* were identified in five participants. The [GRCh38] Xp22.11(23002082-23002458)x1 was detected in one individual (7-0111-003) and the [GRCh38]Xp22.11(23002148-23002522)x1 in four affected brothers (1-0627-003, 1-0627-005, 1-0627-006, and 1-0627-007). These deletions selectively involve a portion of the 3’UTR of the gene and are absent in the DECIPHER database. Overall, the identified *DDX53* variations are predicted to affect protein function through the alteration of conserved residues within functional domains or the loss of a functional transcript/part of the 3’UTR. Additionally, a maternally inherited, 21.8 kb deletion [GRCh38]Xp22.11(22987677-23009507)x1 impacting only *DDX53* and not exons of *PTCHD1-AS* was observed in one affected female. However, this individual also carries a 331 kb *de novo* deletion [GRCh38]7q11.22 (70441029-70771836)x1 impacting 5 exons of *AUTS2* (MIM*607270), a gene strongly associated with ASD^38^. In the aforementioned individual, the *AUTS2* deletion is likely involved in the ASD, but the carrier status of the *DDX53* deletion may also need to be considered in male offspring.

In total, in our extended cohort of 31 individuals, including 28 males and 3 females, we identified 25 distinct *DDX53* variations. These included 18 missense changes, 2 truncating variants, 2 in-frame variants, 2 deletions of the 3’UTR, and one complete gene deletion. Four variants were found in more than one individual, including p.(Glu190Cysfs*3) in two (#3 - SP0039198 and, #4 - SP0042895); p.(Asp575Gly) in two (SP0126624 and 1-0551-003); and p.(Gln591del) in two (SP0066816 and SP0066817). Of note, the [GRCh38]Xp22.11(23002148_23002522)x1 deletion was also identified in four participants from the same family with a similar deletion found in another family with one affected male. Missense changes affected conserved residues within or in proximity to the KH, DEXDc, or HELICc functional domains (Figure 1B). *In silico* tools revealed that these variants are variably deleterious (Table S1 and Table S3) and mostly map to amino acid residues intolerant to variation according to Metadome (Figure 3 and Table S4). One of the in-frame variants included the indel p.(Met381_Leu382delinsIleMet), causing the substitution of two conserved amino acids (GERP scores = 4.3) within the DEXDc domain, and p.(Gln591del), resulting in a deletion of a single conserved residue (GERP score = 4.46) in the C-terminus of the protein. The two truncating variants, which were identified in three participants, featured the stopgain p.(Trp8*) and the frameshift p.(Glu190Cysfs*3). These variants are likely to be deleterious based on the predicted severe impact on protein function and the loss of function intolerance of *DDX53* (probability of loss intolerance (pLI) of 0.66 and a loss of function observed/expected upper bound fraction (LOEUF) of 0.847). Of note, affected females (#4 - SP0042895, SP0003053 and 1-0485-003) either harbored truncating *DDX53* changes or a whole gene deletion. Although further studies are required to elucidate the X-inactivation pattern in these cases, this observation suggests that a severe *DDX53* loss of function may lead to a clinical phenotype in female carriers.

**Figure 3.**
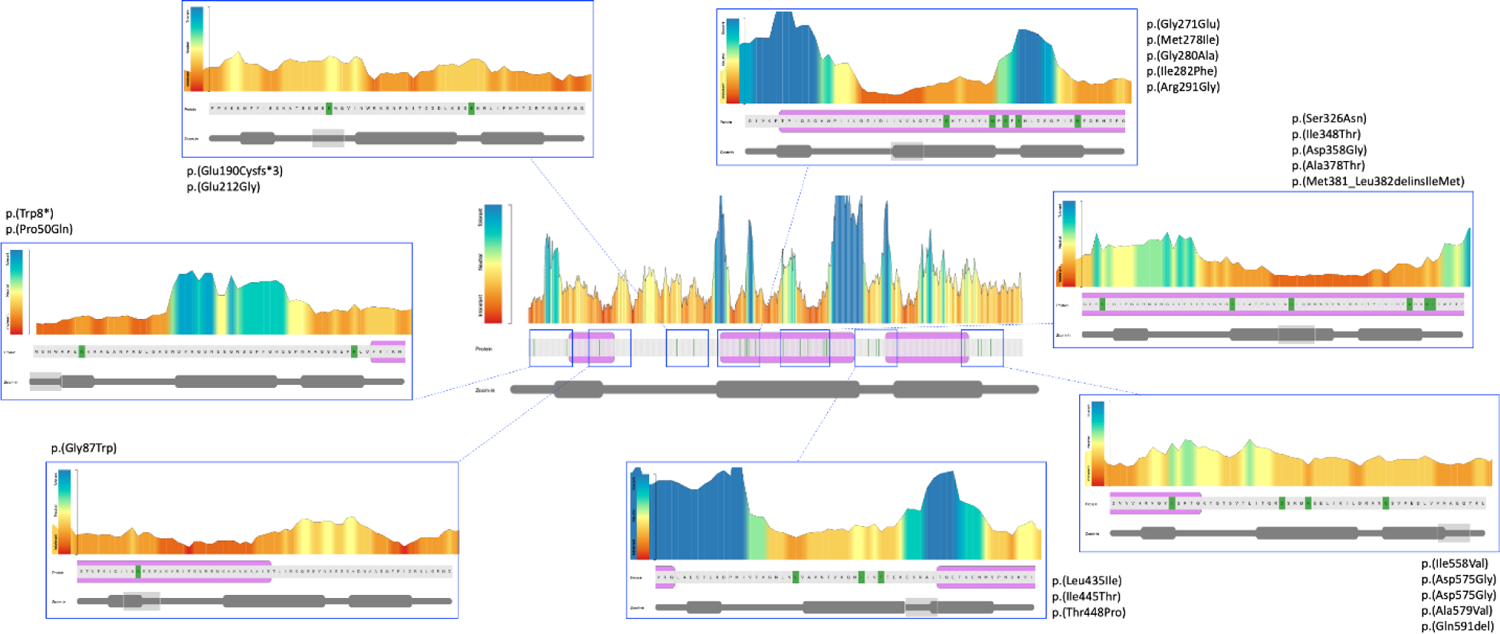
Graphic representation of intolerance to *DDX53* variants. Using the Metadome software (https://stuart.radboudumc.nl/metadome/), we mapped the variants identified in our cohort and in the SFARI and MSSNG datasets to the DDX53 protein. Most variants were found to affect amino acid residues that showed intolerance to variation according to their variation in the gnomAD dataset. While some amino acids were rarely impacted by genetic changes, variants in other residues are absent in gnomAD.

We scrutinized the mouse databases for *Ddx53* and based on the available mouse reference data at UCSC Genome Browser (GRCm39/mm39), there does not appear to be an orthologous transcript in the expected syntenic region (Figure 4)^39^. Instead, there is a gap in alignment between the mouse and human regions; however, that gap does not exist in the DNA of higher mammals.

**Figure 4.**
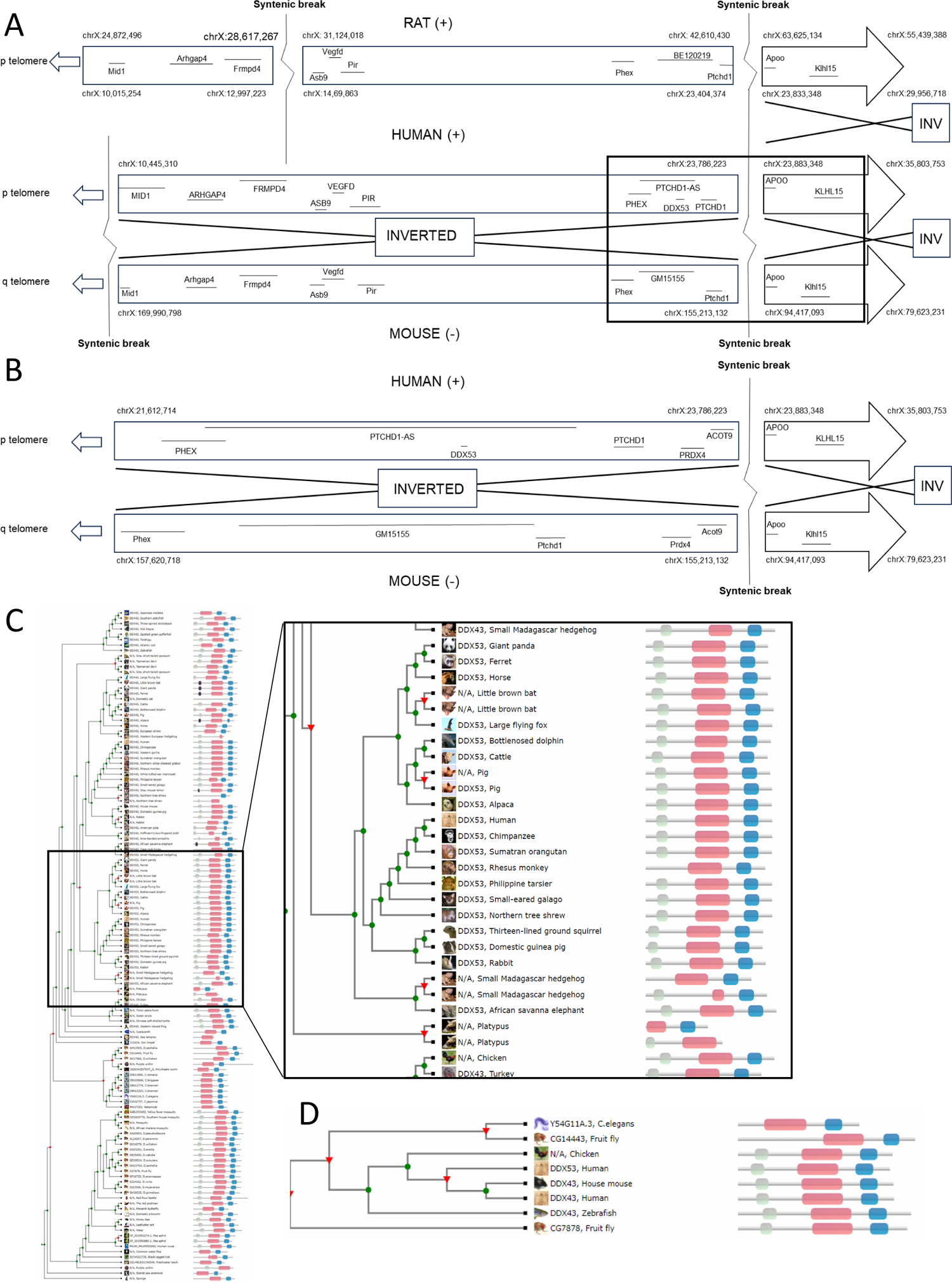
A) Graphical representation of human, mouse, and rat syntenic regions and breaks in chrX:10,445,310-35,803,753 (GRCh38). Syntenic alignment between human and mouse show inverted orientation of two corresponding segments between human and mouse. Syntenic alignment between human and rat is shown to highlight the mouse-specific inversion of this chromosomal segment, and the shared syntenic break located just upstream of the *PTCHD1* gene. B) Inset of human-mouse syntenic region at the *PTCHD1-AS* locus. C) Full phylogenetic tree of *DDX53* and *DDX43* (right panel) created by Treefam. Red triangle denotes a duplication event. Green circle denotes a speciation event. Inset of *DDX53* orthologs (left panel). D) Phylogenetic tree of *DDX53* and *DDX43* in common model organisms.

We designed primers in the repeat-free sequence on each side of the alignment gap to determine if there was any additional sequence in this gap not captured in the murine reference genome.

(forward: ttcaacctatttgtgtgtgcatc; reverse: tccagcacagtattaccagaaatagt). Using BAC clone RP23-405B14 from the C57BL/6 mouse strain as DNA template, PCR was performed with an expected product size of 4,029 bp. Based on gel electrophoresis the product appeared to be ∼4 kb. This product was cloned into a Topo TA cloning vector (Invitrogen), and Sanger sequenced with the original primers, as well as walking primers, to generate the entire 4,029 bp cloned product. The Sanger sequence obtained corresponds with the reference sequence GRCm39/mm39. We repeated this PCR using template DNA from mouse genomic DNA (purchased from Cedarlane) and DNA extracted from internal C57BL/6 mouse brain P14. In each case, the expected product size was amplified and the Sanger sequencing agreed with the reference sequence. All data suggest that there is no additional sequence in this region, in this strain, and therefore a mouse *Ddx53* gene does not appear to be present in the syntenic region to *DDX53*. These observations are further validated because no obvious RNA transcripts were found for *Ddx53* in any database nor in our unpublished murine data. The rat ortholog (*Ddx43*), however, is annotated as a *DDX43/DDX53* homolog and appears to be functional (https://www.genenames.org/tools/hcop/<u>#!/</u>). The naked mole-rat model has a more specific *DDX53* homolog (https://www.ncbi.nlm.nih.gov/gene/101703948/). Other potentially functional homologs are present in rabbits and other small mammals (https://www.ncbi.nlm.nih.gov/gene/168400/ortholog/?scope=9347&term=DDX53) (Figure 4)^40^.

RNA helicases contribute in mRNA metabolism, mediating a range of biochemical activities not restricted to RNA unwinding, such as ribosome biogenesis, RNA degradation, and splicing^41^. While processive RNA helicases function by translocating along the nucleic acids, the non-processive helicases selectively unwind local secondary structures without translocating on their substrates^28; 42^. In humans, the two largest families of helicases are: the DEAD box (DDX) family, encompassing 41 members (https://www.genenames.org/data/genegroup/#!/group/499); and the DEAH box (DHX), including 16 members (https://www.genenames.org/data/genegroup/#!/group/500). The DEAD-box helicases belong to the non-processive group^43^. All DDX helicases share a conserved core consisting of two tandem RecA domains, with recognizable sequence motifs involved in RNA binding, and ATP binding and hydrolysis^41; 42^. Additional C- and N-terminal extensions vary among the different members and consist of auxiliary domains implicated in RNA recognition and the regulation of ATPase activity^41;42^.

Genetic variants affecting the function of DDX helicases have been shown to be involved in other neurodevelopmental disorders^44^. In particular, *de novo* variants in *DDX3X* (MIM * 300160), mapping to chromosome Xp11.3-p11.23, cause a syndromic neural condition (MIM # 300958) with dysmorphism, congenital defects, and neurobehavioral comorbidities including autistic features, either in males or females with different disease severity^45; 46^. More recently, although in absence of supporting functional evidence, missense changes in *DDX54* (MIM * 611665) have been associated with psychomotor development impairment in three unrelated participants, of whom one exhibited repetitive movements and behaviors^44^.

*DDX53* is a little characterized RNA helicase ^21^ whose main functional core consists of the DEAD-like Helicases superfamily (DEXDC) domain, which contains the ATP binding site and mediates the ATP-dependent RNA unwinding activity^21; 47^. Additionally, the protein has two auxiliary domains; the N-terminal K homology (KH) domain, which favors single-strand RNA recognition and binding, and the helicase conserved C-terminal (HELICc) domain, which interacts with the DEXDC domain for the helicase activity^41; 47^. So far, the DDX53 helicase function has only been elucidated in cancer, where its overexpression appears to mediate the acquisition of oncogenic properties in tumoral cells^21; 25; 26^. Based on its elevated gene expression in testis, polymorphisms in *DDX53* may be associated with azoospermia, but this putative finding requires further testing^48; 49^.

It is noteworthy that in our own recent study, Neurogenin 2 (NGN2)-derived cortical excitatory neurons harboring termination codons in *DDX53* showed a comparable activity in the multi-electrode array to isogenic control lines^50^. The lack of a distinctive electrophysiological phenotype and the ability of the knockout induced pluripotent stem cell (iPSC) lines of generating excitatory cortical neurons suggested that loss of *DDX53* does not lead to a glutamatergic dysregulation phenotype, which may argue against an overt role in ASD^50^. However, the isogenic iPSC-derived control neurons investigated in this study showed low levels of *DDX53* transcripts, resulting in protein levels below the threshold for western blot detection^50^. Thus, it is possible that these cells are not the most appropriate model to investigate *DDX53* function; a conjecture further supported through the expression of the *DDX53* gene in the human brain, where significant RNA expression is limited to the cerebellum^15^ (https://www.proteinatlas.org/ENSG00000184735-DDX53/tissue). The cerebellum is responsible for the maturation of nonmotor neural circuitry, thus having a crucial role in guiding cognitive development^51^. Furthermore, specific cerebellar subzones modulate the activity of neocortical substrates contributing to the determination of social interaction patterns during developmentally sensitive periods^51^.

In summary, our genetic data suggest a role for *DDX53* in ASD. Interestingly, at the same Xp22.11 locus, there is also supporting evidence based on non-overlapping microdeletions, for the *PTCHD1-AS* lncRNA, which encompasses *DDX53*, to be independently involved in ASD^15; 20^ (Figure 1). Moreover, at the same locus, for the protein-coding *PTCHD1*, non-synonymous variants have also been found to cause intellectual disability, and in a few individuals, an accompanying diagnosis of autism was present^18^. In murine models, *Ptchd1* deficiency leads to cognitive dysfunction and abnormal behavior with significant hyperactivity; however, despite these behavioral abnormalities they do not strictly resemble the autism spectrum^17; 19^. *Ptchd1-as* cell^20^ and mouse models (unpublished) are being generated. As additional participants with ASD that have sequence-level or deletion variants impacting *DDX53*, *PTCHD1-AS* and/or *PTCHD1* are identified, we anticipate an increasingly complex social-behavioral and clinical presentation when two or more of these co-located genes are impacted. Given the newly found role for *DDX53* in ASD, and that there is no apparent functional ortholog (*Ddx53*) in mouse, generating a ‘humanized’ *Ddx53* murine model may be important for future mouse-modelling of autism.

## Abbreviations

ACMG-AMP: American College of Medical Genetics and Genomics and the Association for Molecular Pathology

ADHD: attention deficit-hyperactivity disorder

ASD: autism spectrum disorder

ES: exome sequencing

iPSC: induced pluripotent stem cell

NMD: nonsense-mediated mRNA decay

SNVs: single nucleotide variants

CNV: copy number variant

SV: structural variant

SFARI: Simons Foundation Autism Research Initiative

EAGLE: evaluating evidence for ASD

## Data Availability

The data that support the findings of this study are available from the corresponding authors upon request or included in the databases mentioned and linked in the manuscript.

https://research.mss.ng

https://www.sfari.org/resource/sfari-gene/

## Acknowledgements

We acknowledge technical support from Bhooma Thiruvahindrapuram, and Sylvia Lamoureux. We thank the research participants and their families (including those participating in MSSNG) as well as the generosity of the donors who support this program. We acknowledge the resources of Autism Speaks and The Centre for Applied Genomics. We are grateful to all of the families described in this paper including those involved in the Autism Speaks MSSNG Project, as well as the SSC and SPARK sites including the principal investigators (A. Beaudet, R. Bernier, J. Constantino, E. Cook, E. Fombonne, D. Geschwind, R. Goin-Kochel, E. Hanson, D. Grice, A. Klin, D. Ledbetter, C. Lord, C. Martin, D. Martin, R. Maxim, J. Miles, O. Ousley, K. Pelphrey, B. Peterson, J. Piggot, C. Saulnier, M. State, W. Stone, J. Sutcliffe, C. Walsh, Z. Warren, and E. Wijsman). We appreciate obtaining access to genetic and phenotypic data on SFARI Base.

## Author Contributions

Conceptualization: MS, SWS; Formal Analysis: MS, JLH, NBS, CS, MSR, JRM, SWS; Funding Acquisition: MS, SFN; SWS; Investigation: MS, CAB, JLH, BT, CS, MSR, JRM, SYK, FZ, SWS; Methodology: MS, SWS; Project Administration: MS, JLH, SWS; Resources: MS, PWF, LG, GA, VP, AB, RK, SP, HFP, LL, PPM, IH, SVM, ED-B, BER, SFN, SWS; Supervision: MS, SWS; Visualization: MS, JLH, CS, MSR, SWS; Writing: MS, JLH, NBS, CS, JRM, SWS

## Web resources

CADD; https://cadd.gs.washington.edu

ClinVar; https://www.ncbi.nlm.nih.gov/clinvar

Combined Annotation Dependent Depletion (CADD); http://cadd.gs.washington.edu

Database of Genomic Variants (DGV); http://dgv.tcag.ca/dgv/app/home

DECIPHER; https://decipher.sanger.ac.uk

Gene Cards; https://www.genecards.org

Gene Matcher; http://www.genematcher.org

Genome Aggregation Database (GnomAD); http://gnomad.broadinstitute.org

Genomic Evolutionary Rate Profiling (GERP); http://mendel.stanford.edu/SidowLab/downloads/gerp/

gnomAD; https://gnomad.broadinstitute.org

Metadome online software; https://stuart.radboudumc.nl/metadome/method

MSSNG; https://research.mss.ng

Online Mendelian Inheritance in Man; https://www.ncbi.nlm.nih.gov/Omim

Protein Atlas; https://www.proteinatlas.org/ENSG00000184735-DDX53/tissue

Polyphen-2; http://genetics.bwh.harvard.edu/pph2/

PubMed; https://www.ncbi.nlm.nih.gov/pubmed

RefSeq; https://www.ncbi.nlm.nih.gov/refseq

SFARI; https://www.sfari.org/resource/sfari-gene/

SIFT; https://sift.bii.a-star.edu.sg

UniProt; https://www.uniprot.org

UCSC Human Genome Database; https://www.genome.ucsc.edu

Varsome; https://varsome.com

Variant Effect Predictor (VEP) - Ensembl; https://www.ensembl.org/info/docs/tools/vep/index.html

---

**Table S1.** Extended *in silico* analysis of *DDX53* variants in our cohort.

**Table S2.** Extended clinical phenotype of participants with *DDX53* variants.

**Table S3.** Extended *in silico* analysis of all *DDX53* variants of participants from SFARI and MSSNG databases.

**Table S4.** Details of intolerance to variation for the amino acid residues affected by *DDX53* variants.

**Table S5.** UDN membership list.

