## Supplementary Material for "Genetic variants in *DDX53* contribute to Autism Spectrum Disorder associated with the Xp22.11 locus"

**1. Supplementary Methods**

**2. Supplementary Results**

**3. Supplementary Tables**

**4. Supplementary References**

### 1. Supplementary Methods

#### *1.1 Participant enrolment*

The participants investigated in the first cohort study were enrolled at several different international research centers and hospitals: Paris-Est Créteil Université, Créteil, France; IRCCS Istituto Giannina Gaslini, Genova, Italy; Adult Autism Centre DSM ASL Città di Torino, 10138 Turin, Italy; Randall Children's Hospital at Legacy Emanuel, Portland, USA; Division of Neurology, Children's Hospital of Philadelphia, USA; Department of Neurology, David Geffen School of Medicine, University of California, Los Angeles, USA; Division of Clinical Genetics, Department of Human Genetics, David Geffen School of Medicine at University of California, Los Angeles, USA.

#### *1.2 Clinical assessment*

For the assessment of the behavioral disorders in the enrolled participants, a thorough neuropsychiatric evaluation was performed in all cases. Additionally, dedicated tests were administrated in some individuals. The Autism Diagnostic Interview-Revised (ADI-R) and Ritvo Autism Asperger Diagnostic Scale–Revised (RAADS–R) were employed in #1 and #2. Autism Diagnostic Schedule (ADOS) and Childhood Autism Rating Scale - Second Edition (CARS-2) were employed for ASD assessment in participant #3. In participant #5, ASD features were investigated through the DSM-V Autistic Spectrum Scales and Capute Developmental Scales.

#### *1.3 Exome sequencing*

After standard DNA extraction, trio-exome sequencing (ES) was performed in participants #1, #2, #6 and genome sequencing in #7, as previously described[1-5]. Quality Control (GC) statistics with FastQC (http://www.bioinformatics.bbsrc.ac.uk/projects/fastqc) was used to assess the quality of the sequence reads. Burrows Wheeler Alignment (BWA) with default parameters was used for reads alignment to the reference human genome (GRCh38 - hg38, UCSC genome assembly). Recalibration of the quality score and for indel realignment and variant calling was performed through the HaplotypeCaller algorithm within the GATK package[6; 7]. Variants were then annotated with ANNOVAR[8] and filtered out for minor allele frequency (MAF) ≤ 0.01 in genomic databases (GnomAD, https://gnomad.broadinstitute.org). Afterwards, *in silico* tools were employed to predict the impact of candidate variants on protein structure and function, including: Combined Annotation Dependent Depletion (CADD, https://cadd.gs.washington.edu), Mutation Taster (http://www.mutationtaster.org), Mutation Assessor (http://mutationassessor.org/r3/), Polyphen-2 (http://genetics.bwh.harvard.edu/pph2/), SIFT (https://sift.bii.a-star.edu.sg), and Splice AI (https://spliceailookup.broadinstitute.org). Sanger sequencing was performed according to standard procedures[5] to confirm the most plausible candidate variants and for parental segregation analysis.

#### *1.4 Copy number variations investigation*

For copy number variant (CNV) assessment, array comparative genomic hybridization (Array-CGH) (Agilent 60K) was performed as previously described in all subjects. Exome-based CNVs calling using a custom-developed analysis tool (XomeAnalyzer) was performed in participants #5 and #6, whereas genome-based CNV calling was also performed in participant #7. Data were filtered and analyzed to identify sequence variants and most deletions and duplications involving three or more coding exons, as previously described[9]. Potential rearrangements were interpreted according to the DECIPHER database (https://decipher.sanger.ac.uk) and Database of Genomic Variants (http://dgv.tcag.ca/dgv/app/home).

### 2. Supplementary Results

*ASD features assessment*

In participant #1, the administration of the ADI-R test led to the following results: qualitative abnormalities of social interaction, score = 17 [cut-off = 10]; qualitative abnormalities of communication skills, score = 12 [cut-off = 10]; repetitive, restricted, and stereotyped behaviors, score = 5 [cut-off = 3]; abnormal development before 36 months, score = 3 [cut-off = 1]. The RAADS-R test showed: language score = 18 [cut-off = 4]; social relatedness score = 92 [cut-off = 31]; sensory–motor score = 18 [cut-off = 16]; circumscribed interests score = 28 [cut-off = 15]

In participant #2, the ADI-R test showed: ADI-R: qualitative abnormalities of social interaction, score = 16 [cut-off = 10]; qualitative abnormalities of communication skills, score = 7 [cut-off = 10]; repetitive, restricted, and stereotyped behaviors, score = 3 [cut-off = 3]; abnormal development before 36 months, score = 1 [cut-off = 1]. The administration of the RAADS-R test showed: language score = 14 [cut-off = 4]; social relatedness score = 61 [cut-off = 31]; sensory–motor score = 21 [cut-off = 16]; circumscribed interests score = 18 [cut-off = 15].

Participant #3 was tested through Preschool Language Scale, where his auditory comprehension score was 50, expressive communication score was 74, and the total language score was 59. He had an autism evaluation done through CARS-2, scoring in the mild to moderate autism spectrum disorder. Additional ASD assessment through the Autism Diagnostic Schedule (ADOS) revealed scores in the autism range and the diagnosis of autism was confirmed using DSM-5 criteria.

In participant #5, the Capute Developmental Scales were administered at 2.5 years, with the following results: overall expressive and receptive language skills at a 12.8-month level (severe deficits); overall visual motor problem solving skills at a 20.7-month level (moderate deficits). On the DSM-V Autistic Spectrum Scales, the participant showed persistent deficits in social communication and social interaction with restricted patterns of behaviors and interests.

Participant #7 was assessed through different tests and scales, all leading to results consistent with an impairment of social interaction and communication skills, confirming a diagnosis of severe autism. Employed tests and scales included Adaptive Behavior Assessment Scale, 3rd Edition (ABAS-3), Beery- Buktenica Visual Motor Integration, 6th Edition (VMI), Developmental Ability Scales, 2nd Edition (DAS-II): Matrices, Picture Similarities, Pattern

Construction, Recall of Digits Forward, Recognition of Pictures, Expressive One Word Picture Vocabulary Test, 4th Edition (EOWPVT-4), NEPSY-II: Comprehension of Instructions, Peabody Picture Vocabulary Test, 5$^{th}$ Edition (PPVT-5), Wechsler Individual Achievement Test, 4th Edition (WIAT-4): Word Reading, Oral Reading Fluency, Reading Comprehension, Spelling, Math Problem Solving, Wechsler Preschool and Primary Scale of Intelligence, 4$^{th}$ Edition (WPPSI-IV), Developmental Ability Scales, 2nd Edition (DAS-II): Verbal Comprehension, NEPSY-II: Memory for Names, Imitating Hand Positions, Block Construction, Woodcock Johnson IV Tests of Achievement: Sentence Reading Fluency, A Qualitative Assessment of Reading Comprehension.

### 3. Supplementary Tables

**Table S1. Extended *in silico* analysis of *DDX53* variants in our cohort.**

**Table S2. Extended clinical phenotype of participants with *DDX53* variants.**

**Table S3. Extended *in silico* analysis of all *DDX53* variants from SFARI and MSSNG databases.**

**Table S4. Details of intolerance to variation for the amino acid residues affected by *DDX53* variants.**

**Table S5. UDN membership list.**
