## Supplementary Table S4 for "Genetic variants in *DDX53* contribute to Autism Spectrum Disorder associated with the Xp22.11 locus"

| **Position** | **Residue** | **Protein domain** | **ClinVar variants at position** | **Related variants** | | **Tolerance**  **score** | **Intolerance status** |
| --- | --- | --- | --- | --- | --- | --- | --- |
|  | | | | **gnomAD*** | **ClinVar*** |  |  |
| **8** | Trp | - | 0 | - | - | 0.39 | Intolerant |
| **50** | Pro | KH | 0 | - | - | 0.64 | Slightly intolerant |
| **87** | Gly | KH | 0 | - | - | 0.38 | Intolerant |
| **190** | Glu | - | 0 | - | - | 0.75 | Neutral |
| **212** | Glu | - | 0 | - | - | 0.69 | Slightly intolerant |
| **271** | Gly | DEXDc | 0 | 24 | 0 | 0.43 | Intolerant |
| **278** | Met | DEXDc | 0 | 32 | 0 | 0.49 | Intolerant |
| **280** | Gly | DEXDc | 0 | 41 | 0 | 1.01 | Slightly tolerant |
| **282** | Ile | DEXDc | 0 | 38 | 0 | 1.57 | Highly tolerant |
| **291** | Arg | DEXDc | 0 | 26 | 0 | 0.46 | Intolerant |
| **326** | Ser | DEXDc | 0 | 30 | 0 | 0.75 | Neutral |
| **348** | Ile | DEXDc | 0 | 28 | 0 | 0.47 | Intolerant |
| **358** | Asp | DEXDc | 0 | 34 | 0 | 0.22 | Intolerant |
| **378** | Ala | DEXDc | 0 | 30 | 0 | 0.47 | Intolerant |
| **381** | Met | DEXDc | 0 | 30 | 0 | 0.63 | Slightly intolerant |
| **382** | Leu | DEXDc | 0 | 35 | 0 | 0.52 | Intolerant |
| **435** | Leu | DEXDc | 0 | - | - | 0.47 | Intolerant |
| **445** | Ile | - | 0 | - | - | 0.71 | Neutral |
| **448** | Thr | - | 0 | - | - | 0.59 | Slightly intolerant |
| **558** | Ile | C2A | 0 | 55 | 0 | 0.91 | Slightly tolerant |
| **575** | Asp | - | 0 | - | - | 0.68 | Slightly intolerant |
| **579** | Ala | - | 0 | - | - | 0.44 | Intolerant |
| **591** | Gln | - | 0 | - | - | 0.53 | Slightly intolerant |

**Table S4. Details of intolerance to variation for the amino acid residues affected by *DDX53* variants.**

Adapted from Metadome (<https://stuart.radboudumc.nl/metadome/dashboard>)
